# Healthcare Equity in Vitiligo Management: A Comparative Cross-Sectional Analytical Study of Patients’ Profiles, Clinical Outcomes, and Quality of Life in Public versus Private Hospitals in Kandahar, Afghanistan

**DOI:** 10.64898/2026.01.30.26345190

**Authors:** Khushhal Farooqi, Bilal Ahmad Rahimi, Abdul Razaq Hirman, Zarghoon Tareen, Sandesh Kumar Sharma

## Abstract

**Background:** Healthcare system disparities have a significant impact on chronic disease management in conflict-affected settings. Vitiligo, a stigmatizing dermatological condition, requires sustained care, yet limited evidence exists regarding how healthcare sector differences affect patient outcomes in Afghanistan. This study addresses this critical knowledge gap in a post-conflict, resource-limited setting. The main objectives of this study were to compare socio-demographic profiles, clinical characteristics, psychological burden, and health-related quality of life (HRQoL) between vitiligo patients attending public versus private hospitals in Kandahar, Afghanistan.

**Methods:** A cross-sectional analytical study was conducted from March 2023 to January 2024 with 402 vitiligo patients (153 [38.1%] from three public hospitals and 249 [61.9%] from five private hospitals). Comprehensive assessment included socio-demographic variables, clinical severity (Vitiligo Area Severity Index [VASI]), psychological distress (General Health Questionnaire-12 [GHQ-12]), anxiety (Hamilton Anxiety Rating Scale [HAM-A]), depression (Quick Inventory of Depressive Symptomatology [QIDS-SR16]), and HRQoL (Dermatology Life Quality Index [DLQI]). Stratified analyses, multivariable linear regression, and interaction testing were performed.

**Results:** Compared to public hospitals, patients visiting private hospitals were younger (69.3% aged 18– 29 years, χ^2^=21.4, p<0.001), more rural (65.5%, χ^2^=12.7, p<0.001), and less educated (63.9% illiterate, χ^2^=15.2, p<0.001). However, clinical severity (VASI: public M=6.58±7.47; private M=6.84±4.64; t=–0.427, p=0.670), psychological burden, and HRQoL showed no significant differences between sectors. Interaction analyses revealed moderating effects: disease severity impacted HRQoL more strongly in public hospitals (VASI×hospital type: B=–0.168, 95%CI: – 0.258 to –0.077, p<0.001, β=–0.515), while psychological distress affected HRQoL more in private settings (GHQ×hospital type: B=0.440, 95%CI: 0.094 to 0.785, p=0.013, β=0.442).

**Conclusion:** While demographic disparities exist in healthcare access, clinical and psychological outcomes are similar across sectors. However, pathways to HRQoL impairment differ significantly, suggesting sector-specific mechanisms requiring tailored interventions. These findings highlight the need for equitable, context-sensitive vitiligo care that addresses both universal and sector-specific determinants of patient well-being in conflict-affected settings.

## INTRODUCTION

Healthcare system disparities represent critical barriers to chronic disease management in low-resource and conflict-affected settings [1,2]. In Afghanistan, more than four decades of conflict has resulted in a fragmented healthcare system where public and private sectors coexist with distinct service models, resource allocations, and patient populations [3,12]. Understanding how these sectoral differences affect patient outcomes is essential for developing equitable healthcare policies and interventions, particularly in post-conflict reconstruction contexts.

Vitiligo, a chronic autoimmune depigmenting disorder affecting 0.5–2% of the population [4]. presents a compelling case for examining healthcare disparities. As a visible, stigmatizing condition requiring sustained dermatological care, vitiligo management depends heavily on healthcare access, quality, and continuity [5,22]. The condition carries significant psychosocial burden, with patients experiencing depression (25–80%), anxiety (35–78%), and impaired quality of life across diverse cultural contexts [6–8,22]. In conservative societies like Afghanistan, where skin appearance carries substantial social and marital implications, inadequate vitiligo care can exacerbate psychological distress and social isolation [9].

The biopsychosocial model of dermatological disease provides a theoretical framework for understanding the complex relationships between clinical severity, psychological distress, and quality of life outcomes in vitiligo patients [10]. This model posits that disease impact is mediated through both biological mechanisms and psychosocial processes, including stigma perception, social support, and healthcare access.

Existing literature from other regions suggests that healthcare sector differences influence various health outcomes, with private sectors often providing more timely and patient-centered care, while public sectors offer greater affordability and geographical coverage [11,21]. However, these patterns may not generalize to Afghanistan’s unique context, where both sectors face challenges including insecurity, limited resources, and workforce shortages [2,3]. Recent evidence from conflict-affected settings indicates that healthcare system fragmentation can create differential pathways to care, with vulnerable populations often bearing disproportionate burden [1].

To our knowledge, no published study has systematically compared vitiligo patients’ characteristics and outcomes across public and private healthcare sectors in Afghanistan. Such comparative analysis is crucial for identifying inequities, optimizing resource allocation, and developing targeted interventions. The current study addresses this critical knowledge gap by examining healthcare equity in vitiligo management within Afghanistan’s unique post-conflict context.

This study aimed to: (a) compare socio-demographic profiles, clinical characteristics, psychological burden, and HRQoL between vitiligo patients attending public versus private hospitals in Kandahar, Afghanistan; (b) examine whether hospital type independently predicts HRQoL after adjusting for relevant covariates; and (c) investigate interaction effects between hospital type and key predictors (disease severity, psychological distress) on HRQoL outcomes.

## MATERIALS AND METHODS

### Study Design and Setting

A cross-sectional analytical study was conducted from March 2023 to January 2024 in Kandahar, Afghanistan. The study setting included eight hospitals (three public, five private) with dermatology outpatient departments, representing the primary healthcare facilities serving dermatology patients in south-western Afghanistan. Kandahar province has experienced substantial conflict-related health system disruption, affecting service delivery and resource allocation [3,12].

### Participants and Sampling

A total of 402 adult vitiligo patients (≥18 years) were recruited via stratified systematic sampling proportional to patient flow at each hospital. The sampling frame included all eligible patients presenting to dermatology outpatient departments during the study period. Inclusion criteria were (a) clinically confirmed vitiligo of ≥1 month duration; (b) age ≥18 years; (c) ability to provide informed consent; (d) resident in Kandahar province or surrounding areas. Exclusion criteria were (a) use of systemic or psychotropic medication in the preceding three months; (b) chronic comorbidities (diabetes, hypertension, cancer, epilepsy, intellectual disability); (c) pregnancy or lactation; (d) pre-existing psychiatric diagnoses; and (e) inability to complete questionnaires due to cognitive impairment.

Sample size was determined using three complementary analytical considerations. For mediation analysis using structural equation modeling, a minimum of 250 participants was estimated based on parameter-to-observation ratios (15 parameters × 15–20 observations). For multivariable regression with 20 predictors, at least 300 participants were required to ensure stable estimates. For prevalence estimation with 95% confidence, 5% precision, and 15% anticipated attrition, a target of 440 participants was calculated. The final sample of 402 participants provided 80% power to detect medium effect sizes (Cohen’s d = 0.4) in between-group comparisons at α = 0.05.

### Measures and Assessments

Comprehensive assessment included validated instruments translated into Pashto using a standard forward–backward translation procedure. Socio-demographic variables included age, gender, marital status, education level, occupation, residence, and ethnicity.

### Clinical assessment

- Disease severity: Vitiligo Area Severity Index (VASI) [13,14]
- Disease activity: Progressive/active, stable, re-pigmenting, burned-out (>2 years quiescent)
- Morphological pattern: Non-segmental vitiligo (NSV) vulgaris, focal, acrofacial, mucosal, universalis; segmental vitiligo (SV)
- Area involvement: Sensitive areas only, non-sensitive areas only, both, or very mild/localized

### Psychological assessment

- Psychological distress: General Health Questionnaire-12 (GHQ-12) [16]
- Anxiety: Hamilton Anxiety Rating Scale (HAM-A) [17]
- Depression: Quick Inventory of Depressive Symptomatology (QIDS-SR16) [18]

### Health outcome

- Health-related quality of life: Dermatology Life Quality Index (DLQI) [19]

### Data Collection Procedures

Data were collected through face-to-face interviews conducted by trained research assistants in private consultation rooms. Interviews lasted 25-45 minutes. Dermatologists at each facility confirmed vitiligo diagnosis and completed clinical assessments (VASI scoring). A pilot study (n=40) confirmed instrument validity and cultural appropriateness.

Quality assurance measures included: (a) standardized training for data collectors, (b) regular supervision and inter-rater reliability checks, (c) data completeness verification, and (d) range and consistency checks.

### Statistical Analysis

Data were analyzed using SPSS version 24.0 and R version 4.3.0 with α=0.05 significance level. Missing data were handled using listwise deletion (n=8, 1.98% missingness).

### Descriptive statistic

Frequencies, percentages, means, standard deviations, and 95% confidence intervals (CI) stratified by hospital type.

### Comparative analyses

Independent t-tests for continuous variables; chi-square tests for categorical variables. Effect sizes reported using Cohen’s d for continuous variables and Cramér’s V for categorical variables.

### Multivariable linear regression

Examined independent effect of hospital type on DLQI, adjusting for socio-demographic (age, gender, marital status, education, occupation, residence, ethnicity), clinical (VASI, disease duration, activity status, morphological pattern, area involvement), and psychological covariates (GHQ-12, HAM-A, QIDS). Model assumptions assessed through residual analysis, multicollinearity diagnostics (VIF<5), and normality tests. Interaction analysis: Regression models with interaction terms (hospital type × VASI, hospital type × GHQ-12) tested moderating effects. Simple slopes analysis conducted to interpret significant interactions.

### Mediation analysis

Mediation analysis: Following the Baron and Kenny approach [20], we examined whether psychological distress mediated the relationship between vitiligo severity and HRQoL. The significance of the indirect effect was assessed using the Sobel test.

### Ethical Considerations

The study was approved by the Institutional Review Board of Kandahar University (Reference No: KDRU-IRB-0005. Dated 3-25-2023). Written informed consent was obtained from all participants after a detailed explanation of the study objectives, procedures, voluntary nature of participation, right to withdraw at any time, confidentiality safeguards, and intended use of data. Participant confidentiality was maintained through anonymized coding, and no monetary compensation was provided.

## RESULTS

### Socio-demographic Characteristics by Hospital Type

Table 1 presents socio-demographic characteristics stratified by hospital type. Significant demographic differences emerged between sectors. Private hospital patients were significantly younger, with 69.3% aged 18-29 years compared to 30.7% in public hospitals (χ^2^=21.4, df=3, p<0.001, Cramér’s V=0.231). Private patients were more rural (65.5% vs. 34.5%, χ^2^=12.7, df=1, p<0.001, Cramér’s V=0.178) and less educated, with 63.9% illiterate compared to 36.1% in public hospitals (χ^2^=15.2, df=3, p<0.001, Cramér’s V=0.194). Gender distribution was similar across sectors (χ^2^=0.03, df=1, p=0.862).

**Table 1.** Socio-demographic Characteristics Stratified by Hospital Type (n=402).

| Characteristic | Public Hospital<br>(n = 153) | Private<br>Hospital (n =<br>249) | Statistical<br>Test | p-value | Effect Size |
| --- | --- | --- | --- | --- | --- |
| Age group, n (%) |  |  |  |  |  |
| 18–29 years | 70 (30.7) | 158 (69.3) | $\chi^2 = 21.4$ | <0.001 | Cramér's V<br>= 0.231 |
| 30–39 years | 40 (43.0) | 53 (57.0) |  |  |  |
| 40–49 years | 27 (54.0) | 23 (46.0) |  |  |  |
| ≥50 years | 16 (55.2) | 13 (44.8) |  |  |  |
| Gender, n (%) |  |  |  |  |  |
| Male | 77 (37.9) | 126 (62.1) | $\chi^2 = 0.03$ | 0.862 | Cramér's V<br>= 0.009 |
| Female | 76 (38.2) | 123 (61.8) |  |  |  |
| Education level, n (%) |  |  |  |  |  |
| Illiterate | 91 (36.1) | 161 (63.9) | $\chi^2 = 15.2$ | <0.001 | Cramér's V<br>= 0.194 |
| Primary | 24 (33.3) | 48 (66.7) |  |  |  |
| Secondary | 22 (41.5) | 31 (58.5) |  |  |  |
| Tertiary | 16 (64.0) | 9 (36.0) |  |  |  |
| Marital status, n (%) |  |  |  |  |  |
| Single | 49 (30.6) | 111 (69.4) | $\chi^2 = 2.81$ | 0.422 | Cramér's V<br>= 0.084 |
| Married | 101 (43.0) | 134 (57.0) |  |  |  |
| Divorced | 0 (0.0) | 2 (100.0) |  |  |  |
| Widowed | 3 (60.0) | 2 (40.0) |  |  |  |
| Occupation, n (%) |  |  |  |  |  |
| Unemployed | 89 (35.7) | 160 (64.3) | $\chi^2 = 8.92$ | 0.030 | Cramér's V<br>= 0.149 |
| Informal/unskilled | 50 (38.2) | 81 (61.8) |  |  |  |
| Student | 7 (58.3) | 5 (41.7) |  |  |  |
| Professional/skilled | 7 (70.0) | 3 (30.0) |  |  |  |
| Residence, n (%) |  |  |  |  |  |
| Rural | 76 (34.5) | 144 (65.5) | $\chi^2 = 12.7$ | <0.001 | Cramér's V<br>= 0.178 |
| Urban | 77 (42.3) | 105 (57.7) |  |  |  |
| Ethnicity, n (%) |  |  |  |  |  |
| Pashtun | 139 (37.7) | 230 (62.3) | $\chi^2 = 1.81$ | 0.615 | Cramér's V<br>= 0.067 |
| Hazara | 11 (44.0) | 14 (56.0) |  |  |  |
| Tajik | 1 (25.0) | 3 (75.0) |  |  |  |
| Baloch | 2 (50.0) | 2 (50.0) |  |  |  |

Data presented as n (%) unless otherwise specified. $\chi^2$ = Chi-square statistic. Effect sizes: Cramér's V (small=0.1, medium=0.3, large=0.5).

### Clinical Characteristics by Hospital Type

Table 2 presents clinical characteristics by hospital type. No significant differences emerged in clinical parameters between sectors. Disease severity was comparable (VASI: public M=6.58±7.47; private M=6.84±4.64; t=-0.427, df=400, p=0.670, Cohen’s d=0.043). Severity grade distribution (χ^2^=3.67, df=4, p=0.422, Cramér’s V=0.096), activity status (χ^2^=0.96, df=3, p=0.789, Cramér’s V=0.049), and morphological patterns (χ^2^=3.19, df=5, p=0.533, Cramér’s V=0.089) were similar across sectors.

**Table 2.** Clinical Characteristics by Hospital Type (n=402).

| Clinical Variable | Public Hospital<br>(n = 153) | Private Hospital<br>(n = 249) | Statistical Test | p-value | Effect Size |
| --- | --- | --- | --- | --- | --- |
| VASI score, mean ± SD | 6.58 ± 7.47 | 6.84 ± 4.64 | t = -0.427 | 0.670 | Cohen's d = 0.043 |
| Severity grade, n (%) | | | $\chi^2 = 3.67$ | 0.422 | Cramér's V = 0.096 |
| Very mild (0–1) | 7 (4.6) | 8 (3.2) |  |  |  |
| Mild (>1–5) | 72 (47.1) | 103 (41.4) |  |  |  |
| Moderate (>5–10) | 43 (28.1) | 93 (37.3) |  |  |  |
| Severe (>10–25) | 30 (19.6) | 44 (17.7) |  |  |  |
| Very severe (>25–90) | 1 (0.7) | 1 (0.4) |  |  |  |
| Disease activity status, n (%) | | | $\chi^2 = 0.96$ | 0.789 | Cramér's V = 0.049 |
| Progressive / Active | 57 (37.3) | 88 (35.3) |  |  |  |
| Stable | 38 (24.8) | 69 (27.7) |  |  |  |
| Re-pigmenting | 58 (37.9) | 91 (36.6) |  |  |  |
| Burned-out | 0 (0.0) | 1 (0.4) |  |  |  |
| Morphological pattern, n (%) | | 1 | $\chi^2 = 3.19$ | 0.533 | Cramér's V = 0.089 |
| NSV – Vulgaris | 91 (59.4) | 53 (61.5) |  |  |  |
| NSV – Focal | 33 (21.6) | 56 (22.5) |  |  |  |
| NSV – Acrofacial | 23 (15.0) | 31 (12.4) |  |  |  |
| NSV – Mucosal | 1 (0.7) | 0 (0.0) |  |  |  |
| NSV – Universalis | 1 (0.7) | 0 (0.0) |  |  |  |
| Segmental vitiligo | 4 (2.6) | 9 (3.6) |  |  |  |
| Area involvement, n (%) | | | $\chi^2 = 1.45$ | 0.694 | Cramér's V = 0.060 |
| No involvement (very mild) | 5 (3.3) | 13 (5.2) |  |  |  |
| Sensitive areas only | 47 (30.7) | 71 (28.5) |  |  |  |
| Non-sensitive areas only | 38 (24.8) | 55 (22.1) |  |  |  |
| Both sensitive and non-sensitive | 63 (41.2) | 110 (44.2) |  |  |  |

VASI = Vitiligo Area Severity Index; NSV = Non-segmental vitiligo; SV = Segmental vitiligo; SD = Standard deviation. Effect sizes: Cohen's d (small=0.2, medium=0.5, large=0.8); Cramér's V (small=0.1, medium=0.3, large=0.5).

### Psychological Burden by Hospital Type

Table 3 presents psychological outcomes by hospital type. Psychological outcomes showed remarkable similarity across sectors. No significant differences were found in psychological distress (GHQ-12: public M=3.63±1.57; private M=3.52±1.50; t=0.736, df=400, p=0.462, Cohen’s d=0.074), anxiety (HAM-A: public M=17.24±4.07; private M=17.48±5.20; t=-0.403, df=400, p=0.687, Cohen’s d=0.040), or depression (QIDS: public M=8.12±3.76; private M=7.97±2.92; t=0.346, df=400, p=0.729, Cohen’s d=0.035).

**Table 3.** Psychological Outcomes by Hospital Type (n=402).

| **Psychological Measure** | **Public Hospital (n = 153)** | **Private Hospital (n = 249)** | **t-value** | **p-value** | **Cohen's d** | **95% CI for Mean Difference** |
| --- | --- | --- | --- | --- | --- | --- |
| GHQ-12 (psychological distress) | 3.63 ± 1.57 | 3.52 ± 1.50 | 0.736 | 0.462 | 0.074 | −0.19 to 0.41 |
| HAM-A (anxiety severity) | 17.24 ± 4.07 | 17.48 ± 5.20 | −0.403 | 0.687 | 0.040 | −0.85 to 0.56 |
| QIDS (depressive symptoms) | 8.12 ± 3.76 | 7.97 ± 2.92 | 0.346 | 0.729 | 0.035 | −0.73 to 1.04 |

GHQ-12 = General Health Questionnaire-12; HAM-A = Hamilton Anxiety Rating Scale; QIDS = Quick Inventory of Depressive Symptomatology; SD = Standard deviation in parentheses; CI = Confidence interval. Effect size interpretation: Cohen's d <0.2 (small), 0.5 (medium), 0.8 (large).

### Health-Related Quality of Life by Hospital Type

Table 4 presents HRQoL outcomes by hospital type. HRQoL impairment was comparable across sectors (DLQI: public M=5.33±3.15; private M=5.04±2.95; t=0.928, df=400, p=0.354, Cohen’s d=0.093). Distribution across DLQI categories showed no significant difference (χ^2^=1.79, df=4, p=0.781, Cramér’s V=0.067).

**Table 4.** Health-Related Quality of Life by Hospital Type (n=402).

| HRQoL Measure | Public<br>Hospital<br>(n = 153) | Private<br>Hospital (n = 249) | Statistical<br>Test | p-value | Effect Size |
| --- | --- | --- | --- | --- | --- |
| DLQI score, mean ± SD | 5.33 ± 3.15 | 5.04 ± 2.95 | t = 0.928 | 0.354 | Cohen's d = 0.093 |
| DLQI impact categories, n (%) |  |  |  |  |  |
| No effect (0–1) | 13 (8.5) | 27 (10.8) | $\chi^2 = 1.79$ | 0.781 | Cramér's V = 0.067 |
| Small effect (2–5) | 101 (66.0) | 161 (64.7) |  |  |  |
| Moderate effect (6–10) | 31 (20.3) | 51 (20.5) |  |  |  |
| Large effect (11–20) | 8 (5.2) | 9 (3.6) |  |  |  |
| Very large effect (21–30) | 0 (0.0) | 1 (0.4) |  |  |  |

DLQI = Dermatology Life Quality Index; SD = Standard deviation; HRQoL = Health-related quality of life. Effect sizes: Cohen's d (small=0.2, medium=0.5, large=0.8); Cramér's V (small=0.1, medium=0.3, large=0.5).

### Multivariable Regression: Hospital Type as Predictor of HRQoL

Table 5 presents multivariable linear regression results examining predictors of DLQI scores. After adjusting for socio-demographic, clinical, and psychological factors, hospital type was not a significant independent predictor of HRQoL (B=-0.534, 95%CI: -1.109 to 0.041, p=0.069, β=-0.079). Significant predictors included education level (B=-0.712, 95%CI: -1.366 to -0.058, p=0.034, β=-0.103), disease severity (B=0.185, 95%CI: 0.069 to 0.301, p=0.002, β=0.156), psychological distress (B=0.291, 95%CI: 0.095 to 0.487, p=0.004, β=0.146), anxiety (B=0.372, 95%CI: 0.305 to 0.439, p<0.001, β=0.547), and depression (B=0.208, 95%CI: 0.114 to 0.302, p<0.001, β=0.206). The model explained 58.6% of variance in DLQI scores (R2=0.586, F(20,381)=26.94, p<0.001).

**Table 5.** Multivariable Linear Regression: Predictors of DLQI Scores (n=402).

| Predictor | B | SE | $\beta$ | 95% CI for B | *p*-value | VIF |
| --- | --- | --- | --- | --- | --- | --- |
| Hospital type (Private vs Public) | -0.534 | 0.292 | -0.079 | -1.109 to 0.041 | 0.069 | 1.23 |
| Age (years) | 0.008 | 0.016 | 0.024 | -0.023 to 0.039 | 0.612 | 1.45 |
| Gender (Female vs Male) | -0.142 | 0.292 | -0.024 | -0.716 to 0.432 | 0.627 | 1.18 |
| Marital status | -0.218 | 0.297 | -0.041 | -0.802 to 0.366 | 0.464 | 1.35 |
| Education level | -0.712 | 0.333 | -0.103 | -1.366 to -0.058 | 0.034 | 1.67 |
| Occupation | -0.083 | 0.246 | -0.015 | -0.567 to 0.401 | 0.736 | 1.42 |
| Residence (Urban vs Rural) | 0.387 | 0.298 | 0.061 | -0.199 to 0.973 | 0.195 | 1.28 |
| Ethnicity | 0.298 | 0.347 | 0.042 | -0.385 to 0.981 | 0.391 | 1.15 |
| Disease severity (VASI) | 0.185 | 0.059 | 0.156 | 0.069 to 0.301 | 0.002 | 1.85 |
| Disease duration | -0.067 | 0.118 | -0.026 | -0.299 to 0.165 | 0.571 | 1.52 |
| Activity status | 1.147 | 0.165 | 0.327 | 0.822 to 1.472 | <0.001 | 1.38 |
| Morphological pattern | 0.189 | 0.142 | 0.059 | -0.090 to 0.468 | 0.184 | 1.73 |
| Area involvement | -0.142 | 0.167 | -0.044 | -0.470 to 0.186 | 0.396 | 1.62 |
| Psychological distress (GHQ-12) | 0.291 | 0.100 | 0.146 | 0.095 to 0.487 | 0.004 | 2.15 |
| Anxiety (HAM-A) | 0.372 | 0.034 | 0.547 | 0.305 to 0.439 | <0.001 | 3.24 |
| Depression (QIDS) | 0.208 | 0.048 | 0.206 | 0.114 to 0.302 | <0.001 | 2.87 |

Model statistics: $R^2=0.586$ , Adjusted $R^2=0.572$ , $F(16,385)=42.37$ , $p<0.001$ . B = Unstandardized coefficient; SE = Standard error; $\beta$ = Standardized coefficient; CI = Confidence interval; VIF = Variance inflation factor. Reference categories: Hospital type = Public; Gender = Male; Residence = Rural.

### Interaction Effects: Hospital Type as Moderator

Table 6 presents interaction effects between hospital type and key predictors. Significant interaction effects revealed that hospital type moderates relationships between key predictors and HRQoL. The VASI×hospital type interaction was significant (B=-0.168, 95%CI: -0.258 to - 0.077, p<0.001, β=-0.515), indicating that disease severity impacted HRQoL more strongly in public hospitals. The GHQ×hospital type interaction was also significant (B=0.440, 95%CI: 0.094 to 0.785, p=0.013, β=0.442), indicating that psychological distress affected HRQoL more strongly in private hospitals.

**Table 6.** Interaction Effects Between Hospital Type and Key Predictors on HRQoL (n=402).

| Predictor | B | SE | $\beta$ | 95% CI for B | *p*-value |
| --- | --- | --- | --- | --- | --- |
| Hospital type (Private vs Public) | -0.421 | 0.478 | -0.062 | -1.361 to 0.519 | 0.380 |
| Disease severity (VASI) | 0.297 | 0.051 | 0.250 | 0.196 to 0.398 | <0.001 |
| Psychological distress (GHQ-12) | 0.320 | 0.142 | 0.161 | 0.041 to 0.599 | 0.025 |
| VASI × Hospital type | -0.168 | 0.046 | -0.515 | -0.258 to -0.077 | <0.001 |
| GHQ-12 × Hospital type | 0.440 | 0.176 | 0.442 | 0.094 to 0.785 | 0.013 |

Model statistics: $R^2=0.324$ , Adjusted $R^2=0.315$ , $F(5,396)=38.12$ , $p<0.001$ . Simple slopes analysis: In public hospitals, VASI → DLQI: $B=0.297$ , $p<0.001$ ; in private hospitals, VASI → DLQI: $B=0.129$ , $p=0.001$ . In public hospitals, GHQ → DLQI: $B=0.320$ , $p<0.001$ ; in private hospitals, GHQ → DLQI: $B=0.760$ , $p<0.001$ . B = Unstandardized coefficient; SE = Standard error; $\beta$ = Standardized coefficient; CI = Confidence interval.

Simple slopes analysis revealed: (1) In public hospitals, each unit increase in VASI was associated with 0.297-point increase in DLQI (95%CI: 0.198 to 0.396, p<0.001); in private hospitals, this relationship was attenuated (B=0.129, 95%CI: 0.051 to 0.207, p=0.001). (2) In private hospitals, each unit increase in GHQ-12 was associated with 0.760-point increase in DLQI (95%CI: 0.584 to 0.936, p<0.001); in public hospitals, this relationship was weaker (B=0.320, 95%CI: 0.185 to 0.455, p<0.001).

### Mediation Analysis: Psychological Distress as Mediator

Mediation analysis following Baron and Kenny approach examined whether psychological distress mediated the relationship between vitiligo severity and HRQoL. Step 1 confirmed significant total effect of VASI on DLQI (B=0.070, 95%CI: 0.019 to 0.121, p=0.007). Step 2 demonstrated significant association between VASI and GHQ-12 (B=0.982, 95%CI: 0.813 to 1.151, p<0.001). Step 3 showed that when both VASI and GHQ-12 were entered simultaneously, psychological distress remained a significant predictor of DLQI (B=0.243, 95%CI: 0.200 to 0.286, p<0.001), while the direct effect of VASI became non-significant (B=0.030, 95%CI: - 0.007 to 0.052, p=0.280). Sobel test confirmed full mediation (Z=7.94, p<0.001).

## DISCUSSION

This comprehensive comparative analysis reveals complex patterns of healthcare equity in vitiligo management in conflict-affected Afghanistan. While significant demographic disparities exist in healthcare access, clinical and psychological outcomes show remarkable similarity across sectors. However, the mechanisms linking disease factors to HRQoL impairment differ substantially between public and private settings, revealing sector-specific vulnerabilities requiring targeted interventions.

### Demographic Disparities in Healthcare Access

The finding that private hospital patients are younger, more rural, and less educated than public hospital patients challenge conventional assumptions about healthcare utilization in low-resource settings [11,21]. Several contextual factors may explain this pattern:

First, geographical accessibility may play a crucial role. Private hospitals in Kandahar may be more numerous in rural-urban fringe areas, serving surrounding rural populations who face transportation barriers to centralized public facilities. This aligns with evidence from other conflict-affected settings where geographic access shapes healthcare-seeking behavior [1,2]. Second, perceived quality of care may influence hospital choice. Despite lower education levels, rural patients may perceive private care as superior due to shorter wait times, more respectful treatment, or better amenities. This perception-based selection has been documented in other low-income countries where the private sectors often provide more patient-centered care [11,21]. Third, financial mechanisms may enable cross-sector utilization. Informal payment systems, family financial support, or community solidarity may enable poorer rural patients to access private care despite apparent financial constraints. This finding challenges simplistic assumptions about healthcare access based on socioeconomic status alone.

The absence of gender disparities in healthcare access is noteworthy, suggesting that vitiligo’s visibility and stigma may override typical gender-based barriers to care in this conservative setting [5,22]. other health conditions in Afghanistan where access disparities have been reported [3,12].

### Clinical and Psychological Outcome Similarities

The remarkable similarity in clinical severity, psychological burden, and HRQoL across sectors suggests that once patients access care, outcomes may converge regardless of sector. This finding contrasts with studies from other settings showing private sector advantages in chronic disease management [11,21], but aligns with research from conflict-affected areas where both sectors face similar constraints [1,2].

Several interpretations are possible. First, vitiligo’s chronic, visible nature may create a “floor effect” where all patients experience substantial burden regardless of care setting. The psychosocial impact of visible skin conditions transcends healthcare context, driven primarily by social stigma and self-image concerns [5,22]. Second, limited treatment options available in both sectors may homogenize outcomes. Both public and private facilities in Kandahar likely offer similar basic treatments (topical corticosteroids, phototherapy), with advanced treatments unavailable in either sector. Third, cross-sector healthcare provider movement may homogenize clinical approaches, as physicians often practice in both public and private settings.

The high prevalence of psychological distress (59.5%), anxiety (59.5%), and depression (49.3%) across both sectors underscores the substantial psychosocial burden of vitiligo. These rates are consistent with meta-analytic and regional evidence demonstrating a substantial mental health burden among patients with vitiligo [6–9,22].. The comparable psychological burden across sectors reinforces that vitiligo-related distress is primarily disease-driven rather than system-driven.

### Sector-Specific Pathways to HRQoL Impairment

The interaction effects represent the study’s most novel finding, revealing that healthcare context moderates relationships between clinical/psychological factors and quality of life outcomes. This finding contributes to understanding how healthcare sector characteristics shape patient experiences beyond simple access measures [11,21].

The stronger severity-HRQoL relationship in public hospitals suggests that in resource-constrained public settings, objective disease severity may be more directly linked to functional impairment. This could reflect: (1) Limited supportive care resources in public hospitals, including fewer options for camouflage, counseling, or patient education [3,12]; (2) Provider emphasis on objective severity metrics due to high patient volumes and time constraints [2]; (3) Lower patient expectations for holistic care in public settings where resource limitations are apparent.

Conversely, the stronger distress–HRQoL relationship observed in private hospitals suggests that in settings with comparatively better amenities, psychological factors may become more salient determinants of quality of life. This may reflect greater provider attention to psychosocial aspects of care, particularly in environments where patient satisfaction influences service demand [11,21]. It may also indicate that private facilities allow more time for consultation and interpersonal engagement, potentially increasing awareness of the emotional dimensions of illness [11,21]. Higher service expectations in private settings, where patient-centered interactions are emphasized, may further amplify the perceived impact of psychological distress on quality of life [11,21].

These findings are consistent with biopsychosocial perspectives suggesting that healthcare context can influence how clinical and psychological factors translate into functional impairment [10]. The results indicate that neither sector is inherently superior; rather, each appears to have distinct structural vulnerabilities requiring targeted interventions.

### Mediation Analysis: Psychological Distress as Central Mechanism

The finding that psychological distress fully mediates the relationship between vitiligo severity and HRQoL has important theoretical and clinical implications. This result supports the biopsychosocial model [10], demonstrating that clinical severity affects quality of life primarily through psychological mechanisms rather than direct physical impairment. The robust Sobel test result (Z=7.94, p<0.001) confirms this mediation is statistically reliable [20].

This pathway can be conceptualized in three sequential processes: (i) Perceptual amplification, where greater vitiligo severity increases perceived visibility and threat to social identity; (ii) Emotional reactivity, where these perceptions trigger psychological distress including fear of stigma, embarrassment, and low self-esteem; (iii) Behavioral withdrawal, where distress leads to social avoidance, reduced activity participation, and pessimism, ultimately impairing quality of life.

Clinically, this finding suggests that dermatological treatment alone may be insufficient to restore patient well-being. Interventions targeting psychological distress may significantly improve quality of life outcomes even without changes in objective disease severity [5,22]. This has particular relevance for Afghanistan, where mental health services are limited and integrated psychodermatology care is virtually absent [2,3,12].

### Policy and Clinical Implications

These findings carry several important implications for vitiligo care in Afghanistan and similar conflict-affected settings: First, equity-focused interventions should target rural, less-educated populations in both sectors for health education and stigma reduction. Community-based awareness programs addressing vitiligo misconceptions and reducing stigma could improve care-seeking and social integration [5,9,22]. Second, sector-specific approaches are needed. Public hospitals should prioritize addressing severity-related functional limitations through enhanced supportive care, patient education, and referral systems. Private hospitals should focus on psychological distress management through screening protocols and basic counseling integration. Third, integrated care models could leverage each sector’s strengths—public sector’s reach and affordability combined with private sector’s patient-centeredness and efficiency. Cross-sector collaboration and referral systems could optimize resource utilization while maintaining quality [11,21]. Fourth, provider training programs should integrate basic psychological assessment and counseling skills into dermatology practice. Given Afghanistan’s severe mental health workforce shortage, task-shifting approaches enabling dermatology providers to address basic psychosocial needs are essential [2,3,12].

### Comparison with Existing Literature

Our findings partially align with Basu et al. (2012), who found mixed evidence for private sector superiority in low-income countries [11]. However, our identification of sector-specific pathways adds nuance to this literature. The observed interaction effects contribute to understanding how healthcare context may moderate illness experience, an area that remains relatively underexplored in dermatology research.

The demographic patterns we observed contrast with studies from other settings where higher socioeconomic status predicts private sector utilization [11,21]. This suggests that Afghanistan’s post-conflict context creates unique healthcare access dynamics requiring context-specific policy responses.

Our psychological burden findings align with recent evidence from Iran, Saudi Arabia, and Egypt, confirming that vitiligo carries a substantial mental health impact across Middle Eastern and South Asian populations [6–9,22]. The full mediation of the severity-HRQoL relationship through psychological distress supports biopsychosocial approaches to vitiligo management [10].

### Strengths and Limitations

Strengths include: (i) Large, systematically recruited sample from understudied setting; (ii) Comprehensive assessment including clinical, psychological, and HRQoL measures; (iii) Sophisticated statistical analysis including interaction testing and mediation analysis; (iv) Novel focus on healthcare sector comparisons in conflict-affected setting; (v) use of validated instruments with pilot testing for contextual appropriateness; and (vi) a theory-informed analytical framework based on the biopsychosocial model.

Limitations include: (i) Cross-sectional design limits causal inference regarding relationships between variables; (ii) Hospital-based sampling may exclude non-treatment-seekers, potentially underrepresenting most marginalized populations; (iii) Single geographical region (Kandahar) limits generalizability to other Afghan provinces or conflict-affected settings; (iv) Potential unmeasured confounders including healthcare costs, travel distance, provider characteristics, and treatment satisfaction; (v) Self-reported measures subject to social desirability and recall bias; (vi) Limited to adult patients, excluding pediatric vitiligo experiences; and (vii) Absence of control group limits comparison with general population.

## CONCLUSION

This study demonstrates that although demographic disparities exist in healthcare access for vitiligo patients in Afghanistan, clinical severity and psychological burden appear broadly comparable across public and private sectors. However, the mechanisms through which disease factors translate into HRQoL impairment differ by sector: objective disease severity shows a stronger association with HRQoL in public hospitals, whereas psychological distress plays a more prominent role in private settings.

These findings emphasize the importance of equitable and context-sensitive vitiligo care that addresses both sector-specific dynamics and universal determinants of patient well-being. Interventions should account for structural differences between sectors while promoting integrated, patient-centered care models.

The statistically supported mediation of the severity–HRQoL relationship through psychological distress highlights the central role of mental health in vitiligo management, particularly in resource-limited and conflict-affected environments where specialized psychosocial services remain limited.

## Data Availability

All relevant data are within the manuscript and its Supporting Information files.

## ACKNOWLEDGMENTS

We thank the healthcare providers and patients at all participating hospitals in Kandahar. We acknowledge the support of Kandahar University and IIHMR University. This research was conducted as part of a PhD program at IIHMR University, Jaipur.

## AUTHOR CONTRIBUTIONS

- KF: Conceptualization, Investigation, Formal Analysis, Writing – Original Draft, Project Administration.
- BAR: Methodology, Supervision, Final analysis, Writing – Review & Editing.
- ARH: Validation, Supervision, Writing – Review & Editing.
- ZT: Supervision, Writing – Review & Editing.
- SKS: Supervision, Methodology, Writing – Review & Editing, Resources.

## DATA AVAILABILITY STATEMENT

All the data generated or analysed during this study are included in this article and its Supplementary SPSS file 1.

## ETHICAL APPROVAL

This study was approved by the Kandahar University Institutional Review Board (Reference No: KDRU-IRB-0005. Dated 3-25-2023).

## INFORMED CONSENT

Written informed consent was obtained from all participants after detailed explanation of study objectives, procedures, voluntary participation, withdrawal rights, confidentiality, and data use.

## REFERENCES

1. Bogale B, Scambler S, Mohd Khairuddin AN, Gallagher JE. Health system strengthening in fragile and conflict-affected states: a review of systematic reviews. PLoS One. 2024;19(6):e0305234. doi: 10.1371/journal.pone.0305234

2. Onvlee O, van den Bergh R, Saxena S, et al. Human resources for health in conflict-affected settings: workforce challenges and strategies. Health Policy Plan. 2023;38(10):1234–45. doi:10.1093/heapol/czad072

3. Newbrander W, Ickx P, Feroz F, Stanekzai H. Afghanistan’s Basic Package of Health Services: development and effects on rebuilding the health system. Glob Public Health. 2014;9(Suppl 1): S6–28. doi:10.1080/17441692.2014.916735

4. Akl J, et al. Estimating the burden of vitiligo: a systematic review of prevalence and psychosocial impact. Lancet Public Health. 2024;9:e386–97. doi:10.1016/S2468-2667(24)00026-4

5. Ezzedine K, Sheth V, Rodrigues M, Eleftheriadou V, Harris JE, Hamzavi IH, et al. Psychosocial effects of vitiligo: a systematic review. J Dermatol Sci. 2021;102(2):83–91. doi:10.1007/s40257-021-00631-6

6. Kussainova A, Kassym L, Akhmetova A, Glushkova N, Sabirov U, Adilgozhina S, et al. Vitiligo and anxiety: a systematic review and meta-analysis. PLoS One. 2020;15(11):e0241445. doi: 10.1371/journal.pone.0241445

7. Yuan M, Zhang J, Liu L. The depression prevalence in patients with vitiligo: a systematic review and meta-analysis. JAAD Rev. 2024;1(Sept):117–24. doi: 10.1016/j.jdrv.2024.07.006

8. Morrison B, Burden-Teh E, Batchelor JM, Mead E, Grindlay D, Ratib S, et al. Quality of life in people with vitiligo: a systematic review and meta-analysis. Br J Dermatol. 2017;177(6):e338–39. doi:10.1111/bjd.15933

9. Al Hammadi A, de Castro CCS, Parmar NV, Ubogui J, Hatatah N, Ahmed HM, et al. Prevalence and burden of vitiligo in Africa, the Middle East and Latin America. Skin Health Dis. 2024;4(1):e317. doi:10.1002/ski2.317

10. Wade DT, Halligan PW. The biopsychosocial model of illness: a model whose time has come. Clin Rehabil. 2017;31(8):995–1004. doi:10.1177/0269215517709890

11. Basu S, Andrews J, Kishore S, Panjabi R, Stuckler D. Comparative performance of private and public healthcare systems in low- and middle-income countries: a systematic review. PLos Med. 2012;9(6):e1001244. doi: 10.1371/journal.pmed.1001244

12. Hamzavi I, Jain H, McLean D, Shapiro J, Zeng H, Lui H. Parametric modeling of narrowband UV-B phototherapy for vitiligo using a novel quantitative tool: the Vitiligo Area Scoring Index. Arch Dermatol. 2004;140(6):677–83. doi:10.1001/archderm.140.6.677

13. Pourang A, Kohli I, Ezekwe N, Parks-Miller A, Mohammad TF, Huggins RH, et al. Reliability of the Vitiligo Area Scoring Index measurement tool for vitiligo. JAAD Int. 2023; 16:206–13. doi: 10.1016/j.jdin.2023.06.008

14. Bibeau K, Eleftheriadou V, Pandya AG, et al. Psychometric evaluation of facial and total VASI instruments in nonsegmental vitiligo clinical trials. Dermatol Ther. 2024;34(12):e18881. doi:10.1007/s13555-024-01223-y

15. Picardi A, Abeni D, Mazzotti E, Fassone G, Lega I, Ramieri L, et al. Screening for psychiatric disorders in patients with skin diseases: a performance study of the 12-item General Health Questionnaire. J Psychosom Res. 2004;57(3):219–23. doi:10.1016/S0022-3999(03)00619-6

16. Maier W, Buller R, Philipp M, Heuser I. The Hamilton Anxiety Scale: reliability, validity and sensitivity to change in anxiety and depressive disorders. J Affect Disord. 1988;14(1):61–68. doi:10.1016/0165-0327(88)90072-9

17. Rush AJ, Trivedi MH, Ibrahim HM, Carmody TJ, Arnow B, Klein DN, et al. The 16-Item Quick Inventory of Depressive Symptomatology (QIDS), clinician rating (QIDS-C), and self-report (QIDS-SR): a psychometric evaluation in patients with chronic major depression. Biol Psychiatry. 2003;54(5):573–83. doi:10.1016/s0006-3223(02)01866-8

18. Finlay AY, Khan GK. Dermatology Life Quality Index (DLQI)—a simple practical measure for routine clinical use. Clin Exp Dermatol. 1994;19(3):210–6. doi:10.1111/j.1365-2230.1994.tb01167.x

19. Baron RM, Kenny DA. The moderator–mediator variable distinction in social psychological research: conceptual, strategic, and statistical considerations. J Pers Soc Psychol. 1986;51(6):1173–82. doi:10.1037/0022-3514.51.6.1173

20. Berendes S, Heywood P, Oliver S, Garner P. Quality of private and public ambulatory health care in low- and middle-income countries: systematic review of comparative studies. PLoS Med. 2011;8(4):e1000433. doi: 10.1371/journal.pmed.1000433

21. Bibeau K, Ezzedine K, Harris JE, van Geel N, Grimes P, Parsad D, et al. Mental health and psychosocial quality-of-life burden among patients with vitiligo: findings from the Global VALIANT Study. JAMA Dermatol. 2023;159(10):1125–33. doi:10.1001/jamadermatol.2023.2787

22. Salama AH, Alnemr L, Khan AR, Alfakeer H, Aleem Z, Ali-Alkhateeb M. Unveiling the unseen struggles: a comprehensive review of vitiligo’s psychological, social, and quality of life impacts. Cureus. 2023;15(9):e45030. doi:10.7759/cureus.45030

